# Age-Dependent Association of Tumor Mutational Burden With Overall Survival in Patients With Melanoma

**DOI:** 10.64898/2026.11.24.26368882

**Authors:** Zhuoyang Lyu, Christine G. Lian, Li Zhou

**Affiliations:** Division of General Internal Medicine and Primary Care, Department of Medicine, Brigham and Women’s Hospital, Boston, Massachusetts; Department of Pathology, Brigham and Women’s Hospital and Harvard Medical School, Boston, Massachusetts; Department of Biomedical Informatics, Harvard Medical School, Boston, Massachusetts

## Abstract

**Importance:** Estimates of the association of tumor mutational burden (TMB) with overall survival in melanoma come largely from pan-cancer subgroups or immunotherapy-restricted cohorts, and whether host factors modify TMB’s prognostic value is unclear.

**Objective:** To determine whether TMB is independently associated with overall survival in a clinically sequenced melanoma cohort, and whether patient age modifies that association.

**Design:** This retrospective, single-institution cohort study ascertained vital status through the National Death Index, Massachusetts death records, and electronic health records, with follow-up through September 7, 2025. Data were analyzed from April to November 2026.

**Setting:** A US academic health system.

**Participants:** Patients with a cancer registry–confirmed melanoma diagnosis whose tumor tissue, collected from January 2012 to May 2024, was sequenced on a clinically implemented targeted next-generation sequencing panel (OncoPanel).

**Exposures:** TMB, analyzed continuously (per doubling) and dichotomized at the 10 mutations/megabase threshold (“TMB-high”).

**Main Outcomes and Measures:** The primary outcome was overall survival from the date of genomic specimen collection (sequencing); secondary outcomes were overall survival from diagnosis and melanoma-specific survival.

**Results:** A total of 1063 patients (635 [59.7%] male; 1027 [96.6%] White; median age at sequencing, 65.6 years [IQR, 56.1-74.0]) were included; 445 (41.9%) died. TMB-high status was associated with better overall survival after adjustment for age, sex, and noncutaneous subtype (hazard ratio [HR], 0.63; 95% CI, 0.51-0.79). Median overall survival was 10.2 vs 4.8 years for TMB-high vs TMB-low patients (log-rank P < .001). Each doubling of TMB was associated with an adjusted HR of 0.80 (95% CI, 0.73-0.87), with no evidence of nonlinearity (P = .31). The association weakened with increasing age (interaction HR per decade, 1.29; 95% CI, 1.11-1.50; P < .001); the adjusted TMB-high HR was 0.46 (95% CI, 0.25-0.83) at age 40 years and 0.84 (95% CI, 0.61-1.17) at 80 years. Immune checkpoint inhibitors (ICIs) were received by 823 (77.4%) patients.

**Conclusions and Relevance:** In this cohort study, TMB from clinically implemented, targeted next-generation sequencing panels was associated with overall survival as a continuous gradient, and the association weakened with increasing age. Because most patients received ICIs, prognostic and predictive contributions could not be separated.

**Key Points:** *Question:* Is tumor mutational burden (TMB) associated with overall survival in melanoma, and does age modify the association?

*Findings:* In this cohort study of 1063 patients with melanoma, high TMB was associated with significantly better overall survival (hazard ratio [HR], 0.63); the association weakened with age (HR, 0.46 at 40 years vs 0.84 at 80 years).

*Meaning:* TMB from clinically implemented targeted next-generation sequencing carries age-dependent prognostic information in melanoma.

## Introduction

Melanoma is among the most lethal common skin cancers, with approximately 112 000 new invasive cases and 8500 deaths estimated in the United States in 2026.^1–3^ Over the past decade, immune checkpoint inhibitors (ICIs) and BRAF/MEK-targeted therapy have substantially improved survival in advanced disease, and targeted next-generation sequencing (NGS) panels are now obtained routinely as part of clinical care.^4–6^ These panels report both the individual somatic driver mutations that define a tumor’s molecular subtype and tumor mutational burden (TMB), a summary measure of somatic mutation density.^7,8^

On the basis of the KEYNOTE-158 trial, the US Food and Drug Administration (FDA) granted accelerated, tissue-agnostic approval of pembrolizumab for previously treated, unresectable or metastatic TMB-high solid tumors, defined as at least 10 mutations per megabase (mut/Mb).^9,10^ Higher TMB has been associated with longer survival after checkpoint blockade both across cancer types and in real-world metastatic melanoma treated with first-line ICI.^11,12^ However, overall survival estimates for melanoma specifically remain scarce, because melanoma is usually a minority subgroup within pan-cancer TMB studies and melanoma-focused analyses have generally been confined to treated or metastatic subsets.^11–13^ Interpretation is further constrained by treatment context: higher TMB has been associated with better survival after ICI but worse survival after other treatments.^13^

Two further gaps remain. First, TMB is rarely modeled alongside the driver mutations identified by the same panel, so whether previously described prognostic associations of these mutations are independent of TMB is unclear.^14,15^ Second, whether host factors, particularly age, modify the prognostic value of TMB has rarely been examined in melanoma. In non–small cell lung cancer, high TMB predicted benefit from ICI in younger but not older patients.^16^ In melanoma, evidence on age and immunotherapy outcomes is mixed.^17,18^ We therefore examined, in a clinically sequenced melanoma cohort, whether TMB was independently associated with overall survival and whether this association varied with patient age.

## Methods

### Design, Setting, and Data Sources

We conducted a retrospective, single-institution cohort study at Mass General Brigham, linking the institutional cancer registry, the electronic health record (EHR), and clinical tumor sequencing on the OncoPanel targeted NGS platform.^6,19^ Deaths were identified from the EHR and through linkage to the National Death Index (NDI),^20^ which covered deaths through December 31, 2020, and to Massachusetts death records (through June 1, 2025), which also served to verify recorded deaths (eMethods in Supplement 1); patients with no evidence of death were censored at last known clinical contact. Specimens were collected between January 2012 and May 2024, and follow-up ended on September 7, 2025, the date of data extraction. The Mass General Brigham Institutional Review Board approved this study (protocol 2026P000862) and waived informed consent because the study was retrospective and posed minimal risk. This study followed the Strengthening the Reporting of Observational Studies in Epidemiology (STROBE) reporting guideline.

### Study Population

Eligible patients met 3 criteria, assessed at the patient rather than the specimen level: (1) a melanoma tumor genomic specimen valid for analysis, meaning tissue-derived and successfully sequenced; (2) a pathology report diagnostic of melanoma; and (3) melanoma confirmed by a dated cancer-registry diagnosis or an International Classification of Diseases, Tenth Revision, Clinical Modification code (C43.x or D03.x) in the EHR. One representative tissue specimen was selected per patient by a prespecified rule (most informative fields, then earliest collection date); liquid biopsies were excluded. Of 1216 eligible patients, we excluded 144 without a cancer-registry melanoma diagnosis dated on or before specimen collection, which was required to ascertain anatomic subtype and diagnosis date, and 9 whose recorded death preceded specimen collection. All analyses were conducted in the remaining 1063 patients (eFigure 1 and eTable 1 in Supplement 1). No age restriction was applied.

**Figure 1.**
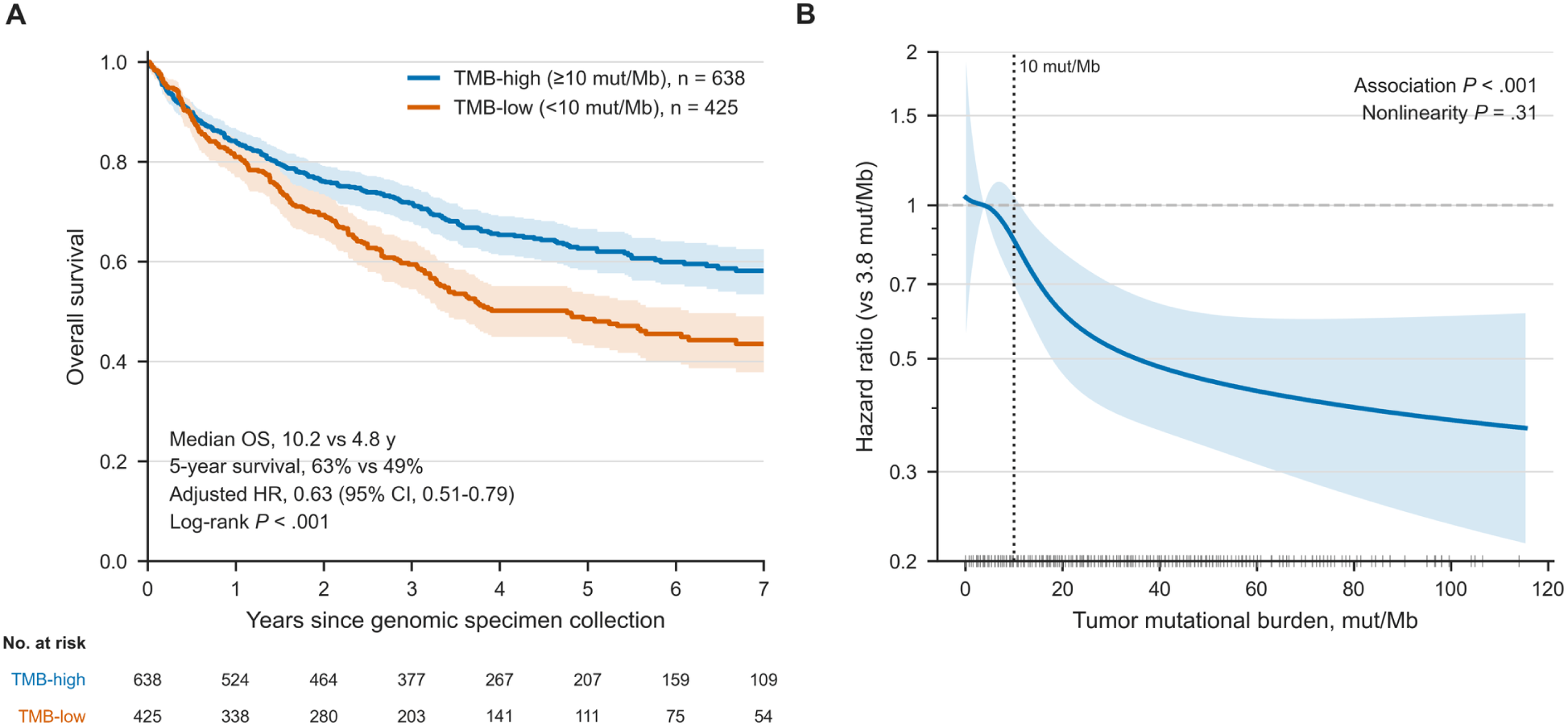
Overall Survival by Tumor Mutational Burden (TMB) A, Kaplan-Meier estimates of overall survival from genomic specimen collection by TMB status (n = 1063; 445 deaths). Shaded areas indicate 95% CIs. The log-rank P value compares the unadjusted curves; the adjusted HR for TMB-high is from the prespecified multivariable model (age at sequencing, sex, and noncutaneous subtype). Curves are drawn to 7 years, the last whole year at which at least 10% of each group remained at risk. B, Restricted cubic spline of the HR for overall survival as a function of TMB, fitted on log(TMB + 1) with knots at 3.0, 9.1, 18.2, and 63.1 mut/Mb and adjusted for age at sequencing, sex, and noncutaneous subtype. HRs are relative to 3.8 mut/Mb (10th percentile); the shaded area indicates the 95% CI. The dotted vertical line marks the 10 mut/Mb threshold. Tick marks along the x-axis indicate observed TMB values. The curve is truncated at the 99th percentile (115 mut/Mb); 11 patients with higher TMB contributed to the fit but are not shown. P values are for the overall association and for nonlinearity on the log scale. HR indicates hazard ratio; mut/Mb, mutations per megabase; OS, overall survival.

### Tumor Mutational Burden and Driver Mutations

TMB (mutations per megabase) was analyzed both continuously and dichotomized at the FDA/KEYNOTE-158 threshold of 10 mut/Mb (“TMB-high”),^9^ with no upper truncation. Because the spline analysis found the burden–hazard relationship to be linear on the logarithmic rather than the arithmetic scale, continuous TMB is reported per doubling; the encodings are compared on model fit in eTable 2 in Supplement 1. Successive OncoPanel versions targeted 275 to 447 genes, so genes added later have lower coverage among earlier-sequenced patients (eMethods in Supplement 1). Mutation status for 11 curated driver genes and The Cancer Genome Atlas (TCGA) molecular class for cutaneous melanoma, assigned hierarchically (*BRAF* V600 > *RAS* > *NF1* > *BRAF* non-V600 > triple wild-type), were derived from the same panel.^8^ *TERT* promoter mutations are undercounted by the coding-region variant filter and *TERT* was excluded from modeling.

### Covariates

Age at specimen collection was used as the age covariate, because it is measured at cohort entry; age at first melanoma diagnosis is reported descriptively for comparability with other melanoma series. Sex and self-reported race and ethnicity were obtained from the EHR. Race categories were American Indian or Alaska Native, Asian, Black or African American, White, other, and unknown; ethnicity was Hispanic or Latino or not Hispanic or Latino. Race and ethnicity were assessed because melanoma incidence and survival differ across these groups and because the composition of the cohort affects the generalizability of its findings.

Tumor anatomic subtype was taken from the cancer registry. Because individual noncutaneous subtypes were uncommon, they were combined into a single indicator of noncutaneous melanoma (any mucosal, ocular, or other noncutaneous site ever recorded in the registry). TCGA molecular class was reported descriptively but not entered into the survival model: it is a 5-level summary of driver mutations measured on the same panel, and median TMB differed more than 6-fold across its levels, so including it alongside TMB would model the exposure twice.

### Outcomes

The primary outcome was overall survival from the date of genomic specimen collection (sequencing), which served as cohort entry. Secondary endpoints were overall survival measured from the date of first melanoma diagnosis, and melanoma-specific and any-cancer-specific survival.

Overall survival was designated the primary endpoint because cause of death was available only from the NDI, which returned a cause for 97.2% of deaths on or before December 7, 2020, and for none thereafter (eTable 3 in Supplement 1). Cause-specific follow-up was therefore censored at that date (eMethods in Supplement 1), which limits these endpoints to approximately the first 4 years of follow-up, so they are reported as supporting analyses. Recurrence-free survival was not analyzed because registry recurrence records do not distinguish a new primary melanoma from progression.

### Statistical Analysis

Unadjusted survival was estimated by the Kaplan-Meier method and compared by the log-rank test.^21^ Associations were estimated by Cox proportional-hazards regression, reporting hazard ratios (HRs) with 95% CIs.^22^ The primary multivariable model contained age at sequencing (per decade), sex, noncutaneous subtype, and TMB. Covariates were chosen a priori from established prognostic factors, not by univariable screening or stepwise selection. Because a covariate’s adjusted HR describes its association holding the others fixed rather than its total effect, only the TMB estimate is interpreted as an exposure–outcome association.^23^ The spline, restricted mean survival time, and delayed-entry analyses used the same covariates. To assess the registry requirement, the age- and sex-adjusted model was refit after adding back eligible patients excluded only for lacking a qualifying registry diagnosis.

Restricted cubic splines of log transformed TMB (log(TMB + 1)) with 4 knots were used to test both for an overall association and for departure from linearity.^24,25^ The proportional-hazards assumption was assessed with scaled Schoenfeld residuals; because hazard ratios may vary over follow-up, this test was not treated as a precondition for the primary analysis, and the 5-year difference in restricted mean survival time is reported as an absolute-scale measure.^26–28^ Cause-specific Cox models censored deaths from other causes, and cumulative incidence treated them as competing events.^29^

We assessed whether the association of TMB with survival differed by each covariate in the multivariable model (age, sex, and noncutaneous subtype); no other interactions were examined. Age was designated in advance as the interaction of primary interest, because TMB’s predictive value has been reported to attenuate with age in non–small cell lung cancer and because immunosenescence offers a candidate mechanism; sex and subtype were examined for completeness and are interpreted as exploratory. Each interaction was fit as a product term within the multivariable model and assessed by likelihood-ratio test, without adjustment for multiple comparisons.

Because misspecifying the main effect of age can produce a spurious TMB × age interaction when TMB and age are correlated, the age interaction was also refit with the main effect of age modeled as a 4-knot restricted cubic spline (eMethods in Supplement 1). To describe the modification across age without categorizing a continuous modifier,^30^ the multivariable model was refit with age modeled as a 4-knot restricted cubic spline in both the main effect and the interaction term, yielding the TMB HR as a continuous function of age with a pointwise 95% CI. A likelihood-ratio test assessed whether the modification departed from linearity (spline vs linear interaction; 2 df).

In a secondary analysis, each of the 11 prespecified driver genes was assessed separately in a model containing age, sex, noncutaneous subtype, TMB, and that gene alone, to address whether a driver mutation carries prognostic information beyond mutational burden. Benjamini-Hochberg q values were computed within the gene family,^31^ and a hazard ratio was reported only when at least 10 deaths occurred in the smaller exposure group.^32^ All tests were 2-sided, and P < .05 (q < .05 for the driver gene analysis) was considered statistically significant. Data were analyzed from April to November 2026 using Python, version 3.12 (Python Software Foundation), with the SciPy (version 1.17.1) and lifelines (version 0.30.3) packages and a fixed random seed of 42.^33,34^

## Results

### Cohort Characteristics

Of 1063 patients with melanoma (median [IQR] age at sequencing, 65.6 [56.1-74.0] years), 428 (40.3%) were female, 635 (59.7%) were male, and 1027 (96.6%) were White. Melanoma was cutaneous in 937 patients (88.1%) and noncutaneous in 126 (11.9%), including 90 (8.5%) with mucosal and 32 (3.0%) with ocular melanoma. Median TMB was 12.2 mut/Mb (IQR, 6.8-25.1) and 638 patients (60.0%) were TMB-high. Compared with the TMB-low group, the TMB-high group was older (median age, 68.5 vs 60.6 years), more often male (425 [66.6%] vs 210 [49.4%]), and less often had noncutaneous melanoma (9 [1.4%] vs 117 [27.5%]) (Table 1). The 153 excluded patients had a similar median age (64.0 years) but were less often TMB-high (78 [51.0%]) (eTable 1 in Supplement 1). At last follow-up, 445 patients (41.9%) had died, with a median follow-up of 4.8 years (IQR, 3.1-7.0) among survivors. Cause of death was unavailable for 240 deaths (53.9%), mostly because they occurred after December 7, 2020 (eTable 3 in Supplement 1).

**Table 1.** Patient Characteristics, Overall and by Tumor Mutational Burden Status.

| **Characteristic** | **Total (N = 1063)** | **TMB-high (n = 638)** | **TMB-low (n = 425)** |
|---|---|---|---|
| Age at sequencing, y |  |  |  |
| Median (IQR) | 65.6 (56.1-74.0) | 68.5 (59.6-76.8) | 60.6 (51.5-70.1) |
| ≤50 | 160 (15.1) | 62 (9.7) | 98 (23.1) |
| 51-70 | 512 (48.2) | 292 (45.8) | 220 (51.8) |
| >70 | 391 (36.8) | 284 (44.5) | 107 (25.2) |
| Age at first melanoma diagnosis, y |  |  |  |
| Median (IQR) | 62 (51-71) | 65 (54-74) | 57 (47-67) |
| Sex |  |  |  |
| Female | 428 (40.3) | 213 (33.4) | 215 (50.6) |
| Male | 635 (59.7) | 425 (66.6) | 210 (49.4) |
| Race<sup>a</sup> |  |  |  |
| American Indian or Alaska Native | 1 (0.1) | 1 (0.2) | 0 (0.0) |
| Asian | 10 (0.9) | 3 (0.5) | 7 (1.6) |
| Black or African American | 3 (0.3) | 0 (0.0) | 3 (0.7) |
| White | 1027 (96.6) | 629 (98.6) | 398 (93.6) |
| Other | 10 (0.9) | 3 (0.5) | 7 (1.6) |
| Unknown | 12 (1.1) | 2 (0.3) | 10 (2.4) |
| Ethnicity<sup>a</sup> |  |  |  |
| Hispanic or Latino | 18 (1.7) | 4 (0.6) | 14 (3.3) |
| Not Hispanic or Latino | 1045 (98.3) | 634 (99.4) | 411 (96.7) |
| Anatomic subtype<sup>b</sup> |  |  |  |
| Cutaneous | 937 (88.1) | 629 (98.6) | 308 (72.5) |
| Mucosal | 90 (8.5) | 6 (0.9) | 84 (19.8) |
| Ocular | 32 (3.0) | 2 (0.3) | 30 (7.1) |
| Other noncutaneous | 4 (0.4) | 1 (0.2) | 3 (0.7) |
| Tumor mutational burden, mut/Mb |  |  |  |
| Median (IQR) | 12.2 (6.8-25.1) | 21.3 (14.4-37.1) | 6.1 (3.8-7.6) |
| Range | 0.0-206.8 | 10.6-206.8 | 0.0-9.9 |
| TCGA molecular class |  |  |  |
| *BRAF* V600 | 288 (27.1) | 138 (21.6) | 150 (35.3) |
| *RAS* | 362 (34.1) | 276 (43.3) | 86 (20.2) |
| *NF1* | 186 (17.5) | 159 (24.9) | 27 (6.4) |
| *BRAF* non-V600 | 27 (2.5) | 21 (3.3) | 6 (1.4) |
| Triple wild-type | 200 (18.8) | 44 (6.9) | 156 (36.7) |
| Immune checkpoint inhibitor exposure |  |  |  |
| Any ICI, any indication | 823 (77.4) | 493 (77.3) | 330 (77.6) |
| Any ICI, melanoma-directed | 726 (68.3) | 451 (70.7) | 275 (64.7) |
| Anti–PD-1 | 801 (75.4) | 481 (75.4) | 320 (75.3) |
| Anti–CTLA-4 | 485 (45.6) | 251 (39.3) | 234 (55.1) |
| Anti–PD-L1 | 27 (2.5) | 8 (1.3) | 19 (4.5) |
| Dual PD-1 and CTLA-4 | 465 (43.7) | 239 (37.5) | 226 (53.2) |
| First ICI relative to sequencing<sup>c</sup> |  |  |  |
| On or before sequencing | 280 (34.0) | 164 (33.3) | 116 (35.2) |
| After sequencing | 543 (66.0) | 329 (66.7) | 214 (64.8) |
| Vital status |  |  |  |
| Deceased | 445 (41.9) | 235 (36.8) | 210 (49.4) |
| Alive (censored) | 618 (58.1) | 403 (63.2) | 215 (50.6) |
| Follow-up among survivors, median (IQR), y | 4.8 (3.1-7.0) | 4.8 (3.2-7.0) | 4.8 (2.9-6.9) |
| Cause of death<sup>d</sup> |  |  |  |
| Melanoma | 160 (36.0) | 85 (36.2) | 75 (35.7) |
| Other cancer | 23 (5.2) | 9 (3.8) | 14 (6.7) |
| Noncancer | 22 (4.9) | 14 (6.0) | 8 (3.8) |
| Not ascertained | 240 (53.9) | 127 (54.0) | 113 (53.8) |

Abbreviations: CTLA-4, cytotoxic T-lymphocyte–associated antigen 4; ICI, immune checkpoint inhibitor; IQR, interquartile range; mut/Mb, mutations per megabase; PD-1, programmed cell death 1; PD-L1, programmed cell death ligand 1; TCGA, The Cancer Genome Atlas; TMB, tumor mutational burden. Data are presented as No. (%) unless otherwise indicated.
<sup>a</sup>Race and ethnicity were self-reported and recorded in the electronic health record; no patient was recorded as Native Hawaiian or Other Pacific Islander or with more than 1 Office of Management and Budget race category. Other indicates race recorded only as some other race: of these 10 patients, 5 reported Hispanic or Latino ethnicity (Cuban, Dominican, Mexican, or Puerto Rican origin) and 5 reported Filipino, Lebanese, Irish, American, or unspecified origin. Two patients recorded as both White and some other race are counted as White. Unknown indicates race recorded as unknown or declined.
<sup>b</sup>Anatomic subtype reflects any site ever recorded; mucosal includes genital, anorectal, sinonasal, and oral sites.
<sup>c</sup>Among patients with any ICI exposure (total, 823; TMB-high, 493; TMB-low, 330).
<sup>d</sup>Among patients who died (total, 445; TMB-high, 235; TMB-low, 210).

### Association of Tumor Mutational Burden With Overall Survival

In the multivariable model, TMB-high status was associated with better overall survival (adjusted HR, 0.63; 95% CI, 0.51-0.79; P < .001; Table 2). Adjusted for age and sex only, the HR was 0.52 (95% CI, 0.43-0.64) and was similar after adding back patients lacking a qualifying registry diagnosis (eTable 2 in Supplement 1).

**Table 2.** Associations of Tumor Mutational Burden and Covariates With Overall Survival (N = 1063; 445 Deaths)

| **Characteristic** | **Unadjusted** |  | **Adjusted, TMB-high model<sup>a</sup>** |  | **Adjusted, continuous TMB model<sup>b</sup>** |  |
|---|---|---|---|---|---|---|
|  | **HR (95% CI)** | ***P* value** | **HR (95% CI)** | ***P* value** | **HR (95% CI)** | ***P* value** |
| Age at sequencing, per decade | 1.30 (1.21-1.40) | <.001 | 1.34 (1.25-1.44) | <.001 | 1.36 (1.26-1.46) | <.001 |
| Sex |  |  |  |  |  |  |
| Female | 1 [Reference] | NA | 1 [Reference] | NA | 1 [Reference] | NA |
| Male | 1.15 (0.95-1.39) | .15 | 1.29 (1.05-1.57) | .01 | 1.31 (1.07-1.59) | .008 |
| Anatomic subtype |  |  |  |  |  |  |
| Cutaneous | 1 [Reference] | NA | 1 [Reference] | NA | 1 [Reference] | NA |
| Noncutaneous | 2.38 (1.88-3.01) | <.001 | 1.90 (1.45-2.49) | <.001 | 1.82 (1.39-2.38) | <.001 |
| TMB status |  |  |  |  |  |  |
| TMB-low (<10 mut/Mb) | 1 [Reference] | NA | 1 [Reference] | NA | NA | NA |
| TMB-high (≥10 mut/Mb) | 0.68 (0.56-0.82) | <.001 | 0.63 (0.51-0.79) | <.001 | NA | NA |
| TMB, per doubling | 0.83 (0.77-0.90) | <.001 | NA | NA | 0.80 (0.73-0.87) | <.001 |
| Harrell C index | NA | NA | 0.636 | NA | 0.644 | NA |

Abbreviations: HR, hazard ratio; mut/Mb, mutations per megabase; NA, not applicable (reference category or variable not in the model); TMB, tumor mutational burden. All estimates are from Cox proportional hazards models in the same 1063 patients (445 deaths); unadjusted HRs are from separate univariable models. Covariates were prespecified on clinical grounds rather than selected by univariable *P* value. Because each adjusted HR is conditional on the other covariates, only the TMB estimate is interpreted as an exposure–outcome association. Model fit under each encoding of TMB is compared in eTable 2 in Supplement 1.
<sup>a</sup>Adjusted for age at sequencing (per decade), sex, noncutaneous subtype, and TMB-high status.
<sup>b</sup>Identical to the TMB-high model except that TMB was entered continuously as log<sub>2</sub>(TMB + 1), with HRs per doubling.

Median overall survival was 10.2 years for TMB-high vs 4.8 years for TMB-low patients, and 5-year survival was 63% vs 49%, respectively (log-rank P < .001) (Figure 1A). Modeled continuously, each doubling of TMB was associated with lower mortality (adjusted HR, 0.80; 95% CI, 0.73-0.87; P < .001); the HR comparing the 75th with the 25th percentile of TMB (25.1 vs 6.8 mut/Mb) was 0.68 (95% CI, 0.58-0.79).

In restricted cubic spline analysis, TMB was associated with overall survival (P < .001 for overall association), with no evidence of departure from linearity on the log scale (P = .31 for nonlinearity; Figure 1B). There was no apparent discontinuity in risk at 10 mut/Mb, and the log-scale encoding fitted better than the 10 mut/Mb dichotomy (eTable 2 in Supplement 1).

There was no evidence of nonproportional hazards for any encoding of TMB (Schoenfeld P ≥ .33). Standardized for age, sex, and subtype, 5-year restricted mean survival time was 0.49 years longer in the TMB-high group (95% CI, 0.22-0.76; eTable 4 in Supplement 1). Associations were in the same direction for overall survival from diagnosis, including with delayed entry at the specimen date, and for melanoma-specific survival (eResults, eTable 4, and eFigures 2 and 3 in Supplement 1), for which each doubling of TMB was associated with an adjusted HR of 0.83 (95% CI, 0.72-0.97).

**Figure 2.**
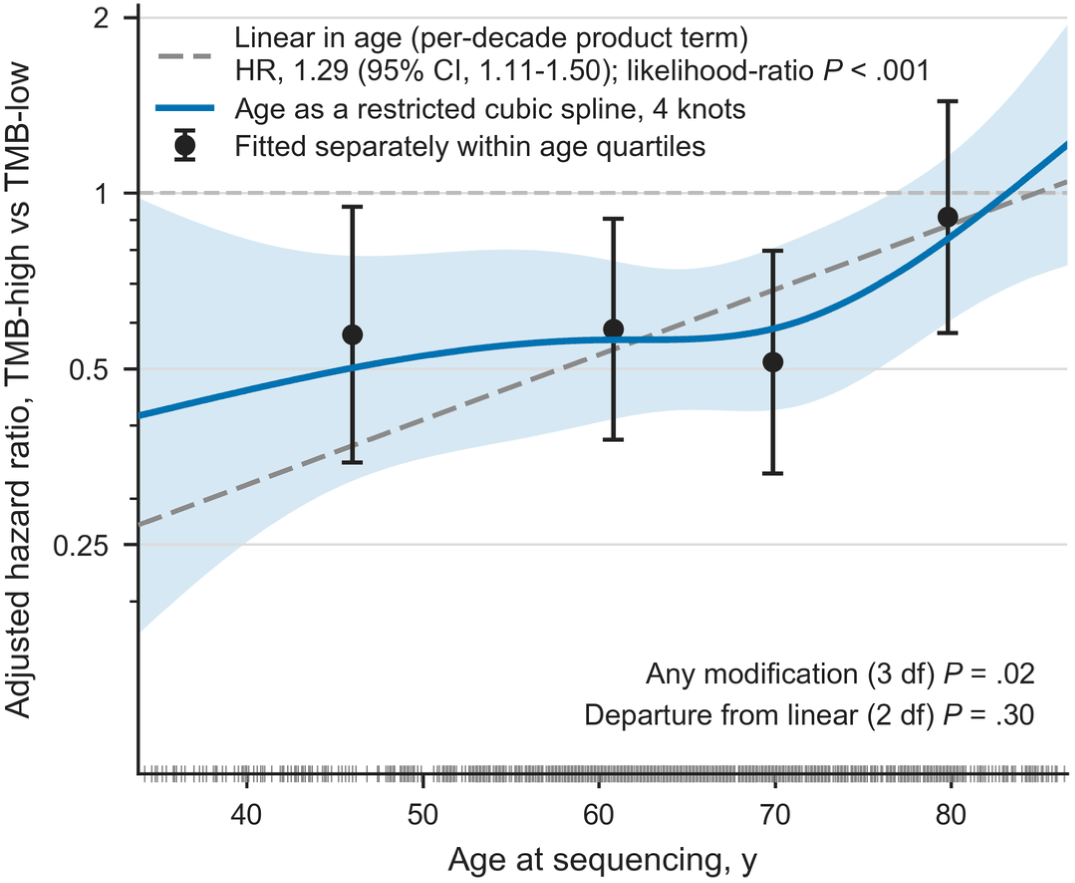
Association of High Tumor Mutational Burden (TMB) With Overall Survival by Age Adjusted hazard ratio for overall survival associated with TMB-high status (≥10 mutations per megabase) as a function of age at sequencing. Estimates are from the prespecified multivariable model (age at sequencing, sex, noncutaneous subtype, and TMB-high; n = 1063; 445 deaths). The solid curve models age as a restricted cubic spline with 4 knots in both the main effect and the interaction; the shaded area indicates the pointwise 95% CI (delta method). The dashed line shows the prespecified linear interaction (product term with age per decade), with its HR and likelihood-ratio P value. P values at lower right are for any modification by age (3 df; spline interaction vs no interaction) and for departure from linearity (2 df; spline vs linear interaction). Circles indicate TMB-high HRs estimated separately within age quartiles, adjusted for sex and noncutaneous subtype; they are not estimates from the plotted model. The curve spans the 5th to 95th percentiles of age (34-87 years), and tick marks along the x-axis indicate observed ages. eFigure 5 in Supplement 1 shows the corresponding analysis for continuous TMB. HR indicates hazard ratio.

In a secondary analysis of 11 driver genes, only *TP53* mutation remained associated with overall survival after false discovery rate correction, and only with TMB dichotomized (HR, 0.70; 95% CI, 0.54-0.89; q = .048; per doubling, q = .22) (eResults and eTables 5 and 6 in Supplement 1).

### Modification of the Tumor Mutational Burden Association by Age

The TMB-high association weakened with increasing age (interaction HR per decade, 1.29; 95% CI, 1.11-1.50; likelihood-ratio P < .001). With age modeled as a restricted cubic spline in both the main effect and the interaction, the adjusted TMB-high HR was 0.46 (95% CI, 0.25-0.83) at age 40 years, 0.56 (95% CI, 0.41-0.77) at 60 years, and 0.84 (95% CI, 0.61-1.17) at 80 years, and the modification did not depart from linearity (P = .30) (Figure 2). The modification was similar with TMB modeled continuously (interaction HR, 1.13 per decade per doubling; 95% CI, 1.06-1.19) and with only the main effect of age modeled as a spline (interaction HR, 1.23 per decade; 95% CI, 1.06-1.44) (eResults, eTable 7, and eFigures 4 and 5 in Supplement 1).

There was no evidence of modification by sex (interaction HR, 0.87; 95% CI, 0.59-1.29) or subtype (interaction HR, 1.08; 95% CI, 0.46-2.55); the subtype estimate was imprecise because few TMB-high patients had noncutaneous melanoma. The age interaction was also present for melanoma-specific survival (interaction HR, 1.43; 95% CI, 1.13-1.81) and for overall survival from diagnosis (eTable 7 in Supplement 1).

### Immune Checkpoint Inhibitor Exposure

Overall, 823 patients (77.4%) received an ICI for any indication and 726 (68.3%) received a melanoma-directed course. ICI exposure was similar in the TMB-high and TMB-low groups for any indication (493 [77.3%] vs 330 [77.6%]) and modestly higher in the TMB-high group for melanoma-directed treatment (451 [70.7%] vs 275 [64.7%]) (Table 1). Most treatment began after cohort entry, however: 543 of the 823 exposed patients (66.0%) received their first course after the specimen date.

## Discussion

In this cohort study of 1063 patients with clinically sequenced melanoma, higher TMB was associated with better overall survival after adjustment for age, sex, and tumor subtype, as a continuous gradient rather than a threshold. This association weakened with increasing age.

These findings are consistent with prior pan-cancer studies and with a real-world cohort of metastatic melanoma treated with first-line immunotherapy.^11,12^ This study adds an overall survival estimate for melanoma from a clinically sequenced population rather than a pan-cancer subgroup, and shows a continuous gradient with no threshold at 10 mut/Mb, which supports reporting continuous TMB alongside the dichotomized cutoff. Both observations should be interpreted in the context of treatment, because the direction of the TMB association differs between patients treated with and without checkpoint inhibitors,^13^ and 77.4% of patients in this cohort received ICIs. Because ICI exposure differed little by TMB status, differential treatment allocation is unlikely to fully explain the association; however, this design cannot distinguish a prognostic association from TMB serving as a marker of immunotherapy responsiveness.

Previously reported prognostic associations of individual driver mutations in melanoma, including *BRAF*, *TP53*, and *NF1*, were generally not adjusted for TMB.^14,35^ In this cohort, only *TP53* mutation remained associated with survival after adjustment for TMB and multiple testing, and only with TMB dichotomized (eTable 5 in Supplement 1), suggesting that some previously reported associations may partly reflect mutational burden. The unfavorable association of *GNAQ*/*GNA11* mutation was attenuated after adjustment for noncutaneous subtype (eTable 6 in Supplement 1); *GNAQ* and *GNA11* mutations are characteristic of uveal melanoma.^36^

To our knowledge, age-dependent modification of the TMB association has not been previously described in melanoma. A parallel pattern has been reported in non–small cell lung cancer treated with ICI,^16^ and immunosenescence is a plausible mechanism. However, several melanoma studies, including a registry study of more than 8000 patients, found ICI efficacy preserved in older patients,^18,37^ and age modifies subtype-specific survival in other cancers outside the immunotherapy context.^38^ This finding should therefore be considered hypothesis generating. If confirmed, TMB from clinically implemented targeted panels could inform survival risk without additional testing, ideally reported as a continuous measure and interpreted in light of patient age; it is not yet a basis for treatment decisions.

### Limitations

This study has limitations. Most importantly, treatment, disease stage, and performance status were not included as covariates, and specimen site (primary tumor vs metastasis) was not captured; because the direction of the association between TMB and survival depends on treatment context, prognostic and predictive contributions could not be separated. This retrospective, single-institution cohort was subject to selection for clinical sequencing, and 96.6% of patients were White, which may limit generalizability. Requiring a qualifying registry diagnosis excluded 144 eligible patients (11.8%); adding back the 137 with valid follow-up did not change the age- and sex-adjusted estimate (eTable 2 in Supplement 1). Only 9 of 638 TMB-high patients had noncutaneous melanoma compared with 117 of 425 TMB-low patients, so subtype adjustment relied on limited overlap between groups, and the estimate was sensitive to it (age- and sex-adjusted HR, 0.52; fully adjusted HR, 0.63). Deaths after December 31, 2020, outside Massachusetts were identified only from the EHR and may be incompletely ascertained, and cause of death was available only through December 7, 2020, which confines the cause-specific endpoints to approximately 4 years. TMB was measured on a targeted panel across successive versions rather than by whole-exome sequencing, and *TERT* promoter mutations were undercounted. Discrimination was modest (C index, 0.64) and calibration was not assessed; these are population-level associations, not a validated individual-risk model.

## Conclusions

In this cohort study of patients with clinically sequenced melanoma, higher tumor mutational burden was associated with better overall survival as a continuous gradient rather than a threshold, and this association weakened with increasing age. Studies with treatment and stage data are needed to separate prognostic from predictive contributions.

## Data Availability

These data are not available for sharing with third parties.

## Article Information

### Author Contributions

Mr Lyu and Dr Zhou had full access to all of the data in the study and take responsibility for the integrity of the data and the accuracy of the data analysis.

*Concept and design:* Lyu, Lian, Zhou. *Acquisition, analysis, or interpretation of data:* Lyu. *Drafting of the manuscript:* Lyu. *Critical review of the manuscript for important intellectual content:* Lian, Zhou. *Statistical analysis:* Lyu. *Obtained funding:* Zhou. *Administrative, technical, or material support:* Zhou. *Supervision:* Lian, Zhou.

### Data Sharing Statement

The patient-level data analyzed in this study were obtained from Mass General Brigham electronic health records, cancer registry, and clinical sequencing records and cannot be shared because of patient privacy protections and institutional restrictions.

### Additional Contributions

The authors used Claude (Anthropic; Claude Opus 5.5 and earlier versions) from October to November 2026 to assist with writing and editing the manuscript. The authors reviewed and edited all content and take full responsibility for the content of this article.

