## Supplement for "Age-Dependent Association of Tumor Mutational Burden With Overall Survival in Patients With Melanoma"

eMethods.

eResults.

eTable 1. Characteristics of the Analytic Cohort and of Excluded Patients.
eTable 2. Sensitivity of the Tumor Mutational Burden Estimate to Covariate Set and to Exposure Encoding.
eTable 3. Verification of Deaths Against Massachusetts Death Records, and Cause-of-Death Ascertainment.
eTable 4. Tumor Mutational Burden Across 4 Survival End Points, and Restricted Mean Survival Time.
eTable 5. Driver-Gene Screen for Overall Survival, and Tumor Mutational Burden by Molecular Class.
eTable 6. Gq-Pathway Mutation and the Triple-Wild-Type Class, Disentangled From Noncutaneous Subtype.
eTable 7. Interaction and Age-Modification Results.
eFigure 1. Cohort Flow Diagram.
eFigure 2. Cumulative Incidence of Melanoma Death by Tumor Mutational Burden Status.
eFigure 3. Delayed-Entry (Left-Truncation) Sensitivity Analysis.
eFigure 4. The Age × TMB-High Interaction Under Alternative Specifications of Age.
eFigure 5. Modification of the Tumor Mutational Burden Association by Age, With Burden Modeled Continuously.
eReferences.

This supplementary material has been provided by the authors to give readers additional information about their work.

### eMethods

#### Covariate selection for the multivariable model

The multivariable model contains age at sequencing, sex, noncutaneous subtype, and tumor mutational burden (TMB). These covariates were specified in advance from established prognostic factors available for the full cohort[1–3]; no variable was added to or removed from the model on the basis of its P value, and no stepwise or univariable-screening procedure was used. eTable 2 reports the TMB estimate under each covariate set examined, so that the sensitivity of the estimate to covariate choice is visible. The Cancer Genome Atlas (TCGA) molecular class was not entered into the model: it is a five-level summary of the same driver mutations that generate mutational burden, and median TMB differs more than sixfold across its levels, so including it alongside TMB would represent the exposure twice rather than adjust for a distinct confounder.[4]

#### Encoding of tumor mutational burden

TMB was modeled in two ways. The dichotomy at 10 mutations per megabase (mut/Mb), which classifies tumors as TMB-high (≥10 mut/Mb) or TMB-low (<10 mut/Mb), is the US Food and Drug Administration threshold based on KEYNOTE-158 and was retained for comparability with the published literature.[5,6] The continuous form is entered as log2(TMB + 1), so that a one-unit increase corresponds to a doubling of burden; the offset accommodates the 2 patients with TMB of 0, and a sensitivity analysis restricted to the 1061 patients with TMB above 0 and using log2(TMB) gave an essentially identical estimate (adjusted hazard ratio, 0.82; 95% CI, 0.76-0.89). The logarithmic form was chosen because the restricted cubic spline found no departure from linearity on the log scale; eTable 2 compares the encodings on a single common sample by partial log-likelihood, Akaike information criterion, Harrell C index, and calibration against the spline.[7]

#### Standardized restricted mean survival time

Because TMB-high patients were older than TMB-low patients and age was strongly associated with mortality, unadjusted restricted mean survival time (RMST) estimates are confounded by that imbalance.[8] We therefore standardized the RMST difference to the cohort's age, sex, and noncutaneous subtype distribution by g-formula direct adjustment. We fit a Cox model with age, sex, and noncutaneous subtype as covariates, stratified by TMB status so that no proportional-hazards constraint was imposed on the TMB term, predicted each patient's survival curve under both TMB assignments, averaged each set of predicted curves across the cohort, and integrated the two averaged curves to each horizon. Confidence intervals were obtained from 1000 bootstrap resamples of the entire procedure.[9] Only the 5-year estimate appears in the main text.

#### Functional form of age in the interaction model

Because a product term can absorb curvature left unmodeled in a main effect,[10] and because TMB-high patients were on average 8 years older than TMB-low patients, the age × TMB-high interaction was refit under alternative specifications of age, all within the multivariable model on the analytic cohort. The association of age with overall survival was mildly nonlinear (restricted cubic spline vs linear, P = .009 with 4 knots and P = .009 with 3 knots), so a linear age term could have biased the interaction estimate. Modeling that curvature with a restricted cubic spline in the main effect of age, while retaining a linear product term, attenuated the interaction from 1.29 to 1.23 (4 knots) and 1.22 (3 knots), and the likelihood ratio test remained statistically significant in both (P = .006 and P = .008). With age entered as a stratification variable by quartile, which gives each quartile its own baseline hazard and so imposes no functional form on age across quartiles but treats age as constant within each quartile, the TMB-high estimate was 0.65 (95% CI, 0.52-0.81). A 1-degree-of-freedom test for linear trend across age quartiles is reported alongside the conventional unstructured heterogeneity test, because the latter ignores the ordering of the quartiles.

#### Displaying the age × TMB interaction across age

Figure 2 in the main text shows the TMB-high hazard ratio as a continuous function of age rather than within age groups, because grouping a continuous modifier discards the variation inside each group and makes the result depend on where the boundaries are placed.[11] The prespecified multivariable model was refit with age entered as a restricted cubic spline with 4 knots, placed at the 5th, 35th, 65th and 95th percentiles of age in the fit sample, the conventional placement for 4 knots,[7] in both its main effect and its product with TMB-high, so that neither the association of age with mortality nor the shape of the modification is forced to a straight line. The hazard ratio at a given age is the exponentiated sum of the TMB-high coefficient and the product block evaluated at that age; its 95% CI is obtained by the delta method from the corresponding submatrix of the covariance matrix and is pointwise, covering the hazard ratio at each single age rather than the whole curve simultaneously. Two nested likelihood-ratio tests are reported, both on the same sample: the spline interaction against the model with no interaction (3 df), which asks whether any modification is present, and the spline interaction against the single per-decade product term (2 df), which asks whether the modification departs from linearity in age. The curve is drawn over the 5th to 95th percentile of observed age so that it does not extrapolate into the sparse tails. As a check on the delta-method interval where the age data are sparse, the curve was refit on 1000 bootstrap resamples of patients with the knots held at their full-sample positions; the percentile intervals agreed closely with the plotted band (at ages 40, 60 and 80 years: 0.20-0.79 vs 0.25-0.83, 0.39-0.77 vs 0.41-0.77, and 0.60-1.18 vs 0.61-1.17). Age quartiles appear in Figure 2 and eFigure 5 only as reference markers; those estimates are fitted separately within each stratum, are not estimates from the plotted model, and are reported in eTable 7.

**Interpretation of multivariable coefficients**

A covariate's adjusted hazard ratio in a jointly fit model describes its association with the outcome holding the other modeled covariates fixed, not that covariate's total or causal effect. Only the TMB estimate is interpreted as an exposure-outcome association; the coefficients for age, sex, and subtype are reported for transparency and are not interpreted causally.

**Panel-version handling**

Successive OncoPanel versions targeted 275 to 447 genes. GNA11 and KRAS were added beginning with version 2.0; patients sequenced only on version 1.0 were not assayed for these 2 genes; their status was treated as unknown rather than wild-type, and they were excluded from analyses involving these genes, giving a denominator of 1031 for those rows in eTable 5. TERT promoter mutations are undercounted because the variant filter retains only coding-region variants, and TERT was excluded from modeling.

**Minimum-events reporting rule**

For a binary covariate, the information about the hazard ratio is carried by the deaths in the smaller exposure group, so the conventional minimum of 10 events per variable, usually applied to whole models, was additionally applied to each binary covariate.[12] Models failing the rule were still fitted, but their estimates are reported as NE (not estimable) in the tables. This rule affected reporting only; no hazard ratio, P value, or q value was changed.

**Immune checkpoint inhibitor ascertainment**

Immune checkpoint inhibitor (ICI) exposure (anti-PD-1, anti-CTLA-4, and anti-PD-L1 agents, alone or in combination) was identified by matching drug names and treatment-plan regimen entries against a prespecified checkpoint-inhibitor list, under two definitions: any ICI (any indication) and a conservative melanoma-directed definition restricted to courses coded to a melanoma diagnosis (International Statistical Classification of Diseases and Related Health Problems, Tenth Revision [ICD-10] code C43.x). ICI exposure was not entered as a Cox covariate because only 280 of 823 ICI-exposed patients (34.0%) began treatment on or before the sequencing date; the median interval from sequencing to first ICI was 13 days (IQR, −16 to 48 days). An ever-received-ICI term would therefore introduce immortal time bias for the majority of exposed patients, who by definition survived long enough after sequencing to start treatment.

**Multiplicity**

Single-gene Cox models were pooled into one Benjamini-Hochberg false-discovery-rate family per endpoint, computed separately within each burden encoding because the 2 adjusted columns of eTable 5 are alternative analyses of the same 11 genes rather than a single family of 22 tests. A q value below .05 was the criterion for statistical significance in the driver-gene analysis; nominal P values are shown for reference. The false discovery rate threshold was .05 throughout, and the random seed was 42.

**Analytic cohort**

Eligibility was assessed at the patient level (eFigure 1). Of 1216 patients meeting the sequencing and diagnostic criteria, 144 had no cancer-registry melanoma diagnosis dated on or before specimen collection (143 had no registry diagnosis and 1 had a registry diagnosis dated after specimen collection) and were excluded, because anatomic subtype, a prespecified covariate, and the date of diagnosis are taken from the registry. A further 9 patients whose recorded date of death preceded specimen collection were excluded as data errors. Every analysis was conducted in the remaining 1063 patients (445 deaths). The one exception is the sensitivity analysis in eTable 2 that adds back the 137 excluded patients with valid follow-up who lacked a qualifying registry diagnosis (n = 1200); because subtype is undefined for most of them, it is adjusted for age and sex only. eTable 1 compares the analytic cohort with the excluded patients.

**Ascertainment of death**

Deaths were identified from 3 sources: the electronic health record (EHR); the National Death Index (NDI), which covered deaths through December 31, 2020; and Massachusetts state death records, which cover deaths occurring in Massachusetts and were available from January 1, 2014, to June 1, 2025 (698 198 records). The state records carry no medical record number and no cause of death, so patients were linked to them by exact match on date of birth, first name, and last name, with the surname at birth also accepted as a last name; only matches to exactly 1 record were used, because looser rules admitted false matches. Of the 1216 eligible patients, 343 matched a single record and none matched more than 1. Censoring was unchanged: patients with no evidence of death were censored at last known clinical contact, and a state record added a death only for a patient recorded as alive whose recorded date of death followed that contact. When sources disagreed on the date of death, the death certificate date took precedence (the NDI date, then the state record date, then the EHR date). Deaths added from the state records have no recorded cause and are counted as cause not ascertained.

eTable 3 compares the state records with the vital status on record before linkage. Of the 425 deaths on record that were dated within the coverage period, 310 (72.9%) were confirmed: 284 of 299 (95.0%) among patients with a Massachusetts home address and 26 of 126 (20.6%) among patients living in other states, whose deaths may have occurred outside Massachusetts. Dates agreed exactly for 307 of the 310 confirmed deaths (99.0%), including all 140 NDI-dated deaths; the other 3 were EHR dates that differed by 1 day and were replaced by the state record date. Among the 622 patients recorded as alive, the state records identified 4 additional deaths (0.6%), 1 of them on or before December 7, 2020, and no state record of death preceded a patient’s last known contact. Of the 1063 patients in the analytic cohort, 689 (64.8%) had a Massachusetts home address. The state records do not cover deaths outside Massachusetts or after June 1, 2025, and the linkage relied on name and date of birth rather than a unique identifier.

**Cause-specific end points**

Cause of death was available only from the National Death Index, which returned a cause for 205 of 211 deaths (97.2%) on or before December 7, 2020, and for none thereafter (eTable 3). Cause-specific follow-up was therefore administratively censored at that date: each patient contributed follow-up from specimen collection to the earlier of last follow-up and December 7, 2020, so the 702 patients sequenced before that date contribute to the cause-specific analyses, and the 6 deaths of unascertained cause within that window were censored at death. Melanoma-specific survival counted melanoma deaths as events, and any-cancer-specific survival counted melanoma and other-cancer deaths. Deaths from other causes were censored in the cause-specific Cox models and treated as competing events in the cumulative incidence analysis (eFigure 2).

### eResults

#### Secondary endpoints

Associations of TMB with survival were in the same direction for all 4 endpoints in eTable 4. Without standardization, 5-year restricted mean survival time was 0.45 years longer in the TMB-high group (95% CI, 0.22-0.68). Each doubling of TMB was associated with lower melanoma-specific mortality (adjusted hazard ratio [HR], 0.83; 95% CI, 0.72-0.97) and any-cancer-specific mortality (adjusted HR, 0.82; 95% CI, 0.72-0.95). The TMB-high estimate was in the same direction but not statistically significant for melanoma-specific death (adjusted HR, 0.82; 95% CI, 0.56-1.18). Cause-specific estimates were less precise than those for overall survival because cause of death was available only for deaths through December 7, 2020, so the cause-specific analyses were based on fewer deaths (eTable 3 and eFigure 2). The age × TMB-high interaction was present for all 3 secondary endpoints (interaction HRs, 1.30-1.43; eTable 7).

#### Driver mutations

After false discovery rate correction, *TP53* was the only driver mutation associated with overall survival independently of TMB, and only with TMB dichotomized (q = .048); with TMB per doubling, no gene met the criterion (minimum q = .06; eTable 5). Median TMB varied more than 6-fold across TCGA molecular classes (eTable 5). Two genes came closest to an independent association and should be considered hypothesis generating. The association of TP53 mutation with better survival was unchanged after adjustment for TMB-high status (unadjusted HR, 0.69; adjusted HR, 0.70; 95% CI, 0.54-0.89; q = .048) but was attenuated with TMB modeled per doubling (adjusted HR, 0.77; 95% CI, 0.59-0.99; q = .22). HRAS mutation (n = 25) was associated with worse survival (adjusted HR, 1.96; 95% CI, 1.17-3.29; q = .06 with either encoding of TMB). NF1 mutation was not associated with survival before or after adjustment. The unfavorable unadjusted associations of GNAQ/GNA11 mutation and of the triple wild-type class were substantially attenuated after adjustment for noncutaneous subtype, although the triple wild-type association persisted within cutaneous melanoma (eTable 6).

#### Sensitivity of the age modification to the modeling of age

Because the TMB-high group was older and the association of age with mortality departed from linearity (P = .009 for nonlinearity), a linear interaction term could partly reflect curvature in the age–mortality relationship rather than a difference between TMB groups. Modeling the main effect of age as a restricted cubic spline attenuated the interaction modestly, from 1.29 to 1.23 per decade (95% CI, 1.06-1.44; likelihood-ratio P = .006) (eTable 7 and eFigure 4). In a Cox model stratified by age quartile, with sex and noncutaneous subtype as the other covariates, the TMB-high HR common to all quartiles was 0.65 (95% CI, 0.52-0.81), and the modification persisted, with a TMB-high HR of 0.54 (95% CI, 0.33-0.87) in the youngest quartile and 0.98 (95% CI, 0.66-1.46) in the oldest (interaction HR, 1.23 per quartile; 95% CI, 1.01-1.48; P = .03 for trend; P = .08 for heterogeneity) (eTable 7). With age modeled as a restricted cubic spline in both the main effect and the interaction, the 95% CI for the TMB-high HR excluded 1 up to approximately age 77 years (Figure 2 in the main text). The modification was also present with TMB modeled per doubling (interaction HR, 1.13 per decade per doubling; 95% CI, 1.06-1.19; likelihood-ratio P < .001), so it did not depend on the 10 mut/Mb cut point (eTable 7 and eFigure 5).

### eTable 1. Characteristics of the Analytic Cohort and of Excluded Patients.

| Characteristic | Analytic cohort (n = 1063) | Excluded (n = 153) |
|---|---|---|
| Age at sequencing, median (IQR), y | 65.6 (56.1-74.0) | 64.0 (54.3-74.7) |
| Male, No. (%) | 635 (59.7) | 80 (52.3) |
| TMB, median (IQR), mut/Mb | 12.2 (6.8-25.1) | 10.6 (6.6-25.1) |
| TMB-high (≥10 mut/Mb), No. (%) | 638 (60.0) | 78 (51.0) |
| Deceased, No. (%) | 445 (41.9) | 67 (43.8) |

*Excluded patients comprise 144 without a cancer-registry melanoma diagnosis dated on or before specimen collection and 9 whose recorded death preceded specimen collection (eFigure 1). Anatomic subtype and diagnosis date are taken from the registry and are unavailable for most excluded patients, so they are not compared. No significance tests are reported. TMB indicates tumor mutational burden.*

### eTable 2. Sensitivity of the Tumor Mutational Burden Estimate to Covariate Set and to Exposure Encoding.

#### A. Covariate set

| Exposure | Covariate set | N | Deaths | HR (95% CI) |
|---|---|---|---|---|
| TMB-high (≥10 mut/Mb) | Unadjusted | 1063 | 445 | 0.68 (0.56-0.82) |
| TMB-high (≥10 mut/Mb) | Age and sex | 1063 | 445 | 0.52 (0.43-0.64) |
| TMB-high (≥10 mut/Mb) | Age and sex, excluded patients added back | 1200 | 499 | 0.53 (0.44-0.64) |
| TMB-high (≥10 mut/Mb) | Age, sex, noncutaneous subtype (multivariable model) | 1063 | 445 | 0.63 (0.51-0.79) |
| TMB, per doubling | Unadjusted | 1063 | 445 | 0.83 (0.77-0.90) |
| TMB, per doubling | Age and sex | 1063 | 445 | 0.74 (0.69-0.81) |
| TMB, per doubling | Age and sex, excluded patients added back | 1200 | 499 | 0.75 (0.70-0.81) |
| TMB, per doubling | Age, sex, noncutaneous subtype (multivariable model) | 1063 | 445 | 0.80 (0.73-0.87) |

*P < .001 for all estimates. All rows were fitted in the analytic cohort (N = 1063; 445 deaths) except the add-back rows, which also include the 137 eligible patients with valid follow-up who were excluded for lacking a registry diagnosis dated on or before sequencing (N = 1200); subtype is undefined for most of these patients, so the add-back rows are adjusted for age and sex only. No variable was added to or removed from the model on the basis of its P value.*

#### B. Model fit by exposure encoding, on one common sample (N = 1063; 445 deaths)

| Encoding of burden | df | Log-likelihood | AIC | Harrell C |
|---|---|---|---|---|
| TMB-high (≥10 mut/Mb) | 1 | -2849.88 | 5707.76 | 0.636 |
| TMB, per 10 mut/Mb (untransformed) | 1 | -2845.79 | 5699.59 | 0.641 |
| TMB, per doubling (log2) | 1 | -2844.82 | 5697.64 | 0.644 |
| Restricted cubic spline on log(TMB+1), 4 knots | 3 | -2843.65 | 5699.30 | 0.645 |

#### C. Calibration against the spline: implied hazard ratio at selected burden values

| TMB, mut/Mb | TMB-high | Per 10 mut/Mb | Per doubling | Spline |
|---|---|---|---|---|
| 10 | 0.63 | 0.92 | 0.76 | 0.85 |
| 20 | 0.63 | 0.81 | 0.62 | 0.61 |
| 40 | 0.63 | 0.63 | 0.50 | 0.48 |
| 100 | 0.63 | 0.29 | 0.37 | 0.38 |

*Abbreviations: AIC, Akaike information criterion; df, degrees of freedom; HR, hazard ratio; mut/Mb, mutations per megabase; TMB, tumor mutational burden.*

*All models were adjusted for age at sequencing, sex and noncutaneous subtype; reference is 3.8 mut/Mb, the 10th percentile of the common sample. The per-doubling form closely approximates the 4-knot spline with a single degree of freedom and has the lowest AIC of any specification, including the spline. The untransformed per-10 term is the poorest-calibrated of the three parametric forms, understating the association where most patients lie and overstating it in the upper tail; it is reported here for completeness only.*

### eTable 3. Verification of Deaths Against Massachusetts Death Records, and Cause-of-Death Ascertainment.

#### A. Deaths on record before linkage (EHR or NDI)

| Group | Deaths, No. | Dated within coverage, No. | Confirmed, No. (%) | Same date of death, No. (%) |
|---|---|---|---|---|
| All deaths on record | 441 | 425 | 310 (72.9) | 307 (99.0) |
| Date of death from the NDI | 206 | 202 | 140 (69.3) | 140 (100) |
| Date of death from the EHR | 232 | 223 | 170 (76.2) | 167 (98.2) |
| Source of date not recorded | 3 | 0 | 0 | NA |
| Massachusetts home address | 306 | 299 | 284 (95.0) | 281 (98.9) |
| Home address in another state | 135 | 126 | 26 (20.6) | 26 (100) |

#### B. Patients recorded as alive before linkage

| Group | Patients, No. | Last contact before June 1, 2025, No. | Death added from state records, No. (%) | State death dated on or before last contact, No. |
|---|---|---|---|---|
| All patients recorded as alive | 622 | 232 | 4 (0.6) | 0 |
| Massachusetts home address | 383 | 97 | 3 (0.8) | 0 |
| Home address in another state | 237 | 133 | 1 (0.4) | 0 |
| Home address missing | 2 | 2 | 0 (0.0) | 0 |

#### C. Cause-of-death ascertainment

| Stratum | Deaths | Cause ascertained | Ascertained, % | Melanoma | Other cancer | Noncancer |
|---|---|---|---|---|---|---|
| **By calendar date of death, relative to the linkage cutoff** | | | | | | |
| On or before December 7, 2020 | 211 | 205 | 97.2 | 160 | 23 | 22 |
| After December 7, 2020 | 234 | 0 | 0.0 | 0 | 0 | 0 |
| **By time from sequencing to death** | | | | | | |
| 0-1 y | 180 | 106 | 58.9 | 89 | 10 | 7 |
| 1-2 y | 98 | 55 | 56.1 | 43 | 6 | 6 |
| 2-3 y | 63 | 26 | 41.3 | 18 | 4 | 4 |
| 3-4 y | 59 | 15 | 25.4 | 9 | 2 | 4 |
| 4-5 y | 14 | 3 | 21.4 | 1 | 1 | 1 |
| 5-7 y | 21 | 0 | 0.0 | 0 | 0 | 0 |
| ≥7 y | 10 | 0 | 0.0 | 0 | 0 | 0 |

*Panels A and B compare Massachusetts state death records (deaths occurring in Massachusetts, January 1, 2014, to June 1, 2025) with vital status recorded before linkage. A death was confirmed when a single state record matched on date of birth, first name, and last name; same-date percentages are among confirmed deaths. The 3 discordant dates were EHR dates 1 day from the state record date and were replaced by it. Home address is from the EHR. EHR indicates electronic health record; NA, not applicable; NDI, National Death Index.*

*In panel C, both blocks include all 445 deaths in the analytic cohort. Cause of death was available only from the National Death Index (NDI), which covered deaths through December 31, 2020; December 7, 2020, was the last date of death for which the NDI returned a cause. Deaths after December 31, 2020, were identified from the electronic health record and Massachusetts state death records, neither of which provided a cause. Ascertainment is therefore a step function of calendar date. The apparent decline with follow-up time in the second block is an artifact of that step, not a property of follow-up itself: long follow-up is observable only for patients sequenced early enough that their late deaths still precede the cutoff. Cause-specific analyses (eTables 4 and 7; eFigure 2) therefore censor follow-up at December 7, 2020 (eMethods). Under the multivariable model, the cause-specific HR for TMB-high was 0.82 (95% CI, 0.56-1.18; n = 702) with this administrative censoring, 0.77 (0.53-1.13; n = 823) when patients whose cause of death could not be ascertained were excluded instead, and 0.85 (0.58-1.24; n = 1063) when those deaths were censored at death without censoring follow-up at the cutoff, so the estimate did not depend on this choice.*

### eTable 4. Tumor Mutational Burden Across 4 Survival End Points, and Restricted Mean Survival Time.

| Endpoint | Exposure | N (deaths) | Unadjusted HR (95% CI) | Adjusted HR (95% CI) |
|---|---|---|---|---|
| Overall survival, from sequencing (primary) | TMB-high | 1063 (445) | 0.68 (0.56-0.82) | 0.63 (0.51-0.79) |
| Overall survival, from sequencing (primary) | Per doubling | 1063 (445) | 0.83 (0.77-0.90) | 0.80 (0.73-0.87) |
| Overall survival, from first melanoma diagnosis | TMB-high | 1063 (445) | 0.67 (0.55-0.80) | 0.65 (0.52-0.80) |
| Overall survival, from first melanoma diagnosis | Per doubling | 1063 (445) | 0.84 (0.78-0.91) | 0.83 (0.76-0.90) |
| Melanoma-specific survival | TMB-high | 702 (160) | 0.75 (0.55-1.02) | 0.82 (0.56-1.18) |
| Melanoma-specific survival | Per doubling | 702 (160) | 0.83 (0.72-0.94) | 0.83 (0.72-0.97) |
| Any-cancer-specific survival | TMB-high | 702 (183) | 0.70 (0.52-0.93) | 0.73 (0.52-1.03) |
| Any-cancer-specific survival | Per doubling | 702 (183) | 0.82 (0.72-0.93) | 0.82 (0.72-0.95) |

**Restricted mean survival time difference on the primary endpoint (TMB-high minus TMB-low)**

| Standardization | Horizon | Difference in life-years (95% CI) |
|---|---|---|
| Unadjusted | 1 y | +0.01 (-0.02 to +0.04) |
| Unadjusted | 3 y | +0.16 (+0.05 to +0.29) |
| Unadjusted | 5 y | +0.45 (+0.22 to +0.68) |
| Standardized for age, sex, and subtype | 1 y | +0.01 (-0.02 to +0.05) |
| Standardized for age, sex, and subtype | 3 y | +0.18 (+0.03 to +0.33) |
| Standardized for age, sex, and subtype | 5 y | +0.49 (+0.22 to +0.76) |

*Abbreviations: HR, hazard ratio; TMB, tumor mutational burden.*

*Adjusted estimates are from the prespecified multivariable model (age at sequencing, sex, noncutaneous subtype). Overall survival end points include all 1063 patients; cause-specific end points are administratively censored at December 7, 2020 (eTable 3), so they include the 702 patients sequenced before that date. Restricted mean survival time differences are shown with bootstrap 95% CIs (1000 resamples); because TMB-high patients were older and older age was strongly associated with mortality, the unadjusted difference is confounded by age and biased toward the null, and the standardized rows apply g-formula direct adjustment to the cohort's own covariate distribution. The 5-year standardized value is the one quoted in the main text.*

### eTable 5. Driver-Gene Screen for Overall Survival, and Tumor Mutational Burden by Molecular Class.

#### A. Driver-gene screen

| Gene | Mutated, n (deaths) | Unadjusted HR (95% CI) | Multivariable model + gene, burden as TMB-high | Multivariable model + gene, burden per doubling |
|---|---|---|---|---|
| BRAF | 414 (142) | 0.66 (0.54-0.81); P<.001 | 0.86 (0.70-1.06); P=.17; q=.31 | 0.88 (0.71-1.09); P=.23; q=.42 |
| NRAS | 317 (136) | 1.06 (0.87-1.30); P=.56 | 1.16 (0.94-1.42); P=.17; q=.31 | 1.10 (0.89-1.35); P=.37; q=.51 |
| KIT | 85 (33) | 0.83 (0.58-1.18); P=.29 | 0.69 (0.48-0.98); P=.04; q=.12 | 0.76 (0.53-1.09); P=.13; q=.36 |
| NF1 | 311 (121) | 0.90 (0.73-1.11); P=.34 | 0.89 (0.71-1.11); P=.29; q=.40 | 1.05 (0.82-1.35); P=.68; q=.70 |
| TP53 | 251 (83) | 0.69 (0.54-0.88); P=.002 | 0.70 (0.54-0.89); P=.004; q=.048 | 0.77 (0.59-0.99); P=.04; q=.22 |
| CDKN2A | 204 (75) | 0.85 (0.67-1.09); P=.21 | 1.03 (0.80-1.33); P=.83; q=.83 | 1.10 (0.85-1.43); P=.47; q=.57 |
| PTEN | 82 (36) | 1.09 (0.78-1.53); P=.62 | 1.22 (0.86-1.72); P=.26; q=.40 | 1.28 (0.90-1.80); P=.16; q=.36 |
| GNAQ | 32 (20) | 2.23 (1.42-3.49); P<.001 | 1.61 (1.02-2.56); P=.04; q=.12 | 1.55 (0.98-2.46); P=.06; q=.23 |
| GNA11 | 40 (21) | 1.48 (0.95-2.29); P=.08 | 1.19 (0.76-1.87); P=.44; q=.53 | 1.28 (0.82-2.02); P=.28; q=.44 |
| KRAS | 32 (11) | 0.80 (0.44-1.46); P=.48 | 0.81 (0.44-1.48); P=.49; q=.54 | 0.89 (0.48-1.63); P=.70; q=.70 |
| HRAS | 25 (15) | 1.69 (1.01-2.83); P=.046 | 1.96 (1.17-3.29); P=.01; q=.06 | 2.11 (1.25-3.54); P=.005; q=.06 |

*Each gene was added, one at a time, to the prespecified multivariable model, so every adjusted estimate is conditioned on age at sequencing, sex, noncutaneous subtype and mutational burden. Because burden is a term in every model, each gene estimate is adjusted for burden by construction and indicates whether a driver mutation carries prognostic information beyond that captured by burden. All models for a gene are fitted on one common complete-case sample (n = 1063; n = 1031 for GNA11 and KRAS, which entered the panel at version 2.0), so differences between the unadjusted and adjusted columns reflect adjustment rather than a change of sample. The q value is the Benjamini-Hochberg false discovery rate within the 11-gene family, computed separately within each encoding.*

#### B. Tumor mutational burden by TCGA molecular class

| TCGA molecular class | n | Median TMB, mut/Mb [IQR] |
|---|---|---|
| NF1 | 186 | 35.4 [19.8-51.5] |
| RAS | 362 | 16.0 [10.6-26.6] |
| BRAF non-V600 | 27 | 16.0 [11.0-30.6] |
| BRAF V600 | 288 | 9.9 [6.8-13.7] |
| Triple wild-type | 200 | 5.3 [3.8-8.5] |

*Abbreviations: HR, hazard ratio; IQR, interquartile range; mut/Mb, mutations per megabase; TCGA, The Cancer Genome Atlas; TMB, tumor mutational burden.*

*N = 1063 (TMB available for all patients). Classes are mutually exclusive and hierarchically assigned (BRAF V600 > RAS > NF1 > BRAF non-V600 > triple wild-type). Median burden differs 6.6-fold between the highest and lowest class (omnibus Kruskal-Wallis P < .001), which is one reason driver-gene estimates can change after adjustment. This panel is descriptive and is not a survival analysis.*

### eTable 6. Gq-Pathway Mutation and the Triple-Wild-Type Class, Disentangled From Noncutaneous Subtype.

| Model | Covariate | N (deaths) | HR (95% CI) | P |
|---|---|---|---|---|
| Univariable | GNAQ/GNA11 (Gq pathway) | 1063 (445) | 1.80 (1.31-2.48) | <.001 |
| Univariable | Triple-wild-type class | 1063 (445) | 1.68 (1.35-2.07) | <.001 |
| Univariable | Noncutaneous subtype | 1063 (445) | 2.38 (1.88-3.01) | <.001 |
| Joint, mutually adjusted (+ age, sex) | GNAQ/GNA11 (Gq pathway) | 1063 (445) | 1.28 (0.91-1.79) | .15 |
| Joint, mutually adjusted (+ age, sex) | Triple-wild-type class | 1063 (445) | 1.29 (1.01-1.67) | .045 |
| Joint, mutually adjusted (+ age, sex) | Noncutaneous subtype | 1063 (445) | 2.04 (1.53-2.72) | <.001 |
| Cutaneous melanoma only (+ age, sex) | GNAQ/GNA11 (Gq pathway) | 937 (358) | 1.07 (0.65-1.74) | .79 |
| Cutaneous melanoma only (+ age, sex) | Triple-wild-type class | 937 (358) | 1.41 (1.06-1.89) | .02 |
| Cutaneous melanoma only (+ age, sex) | GNAQ mutation alone | 937 (358) | NE | — |

*Abbreviations: HR, hazard ratio; NE, not estimable (fewer than 10 deaths in the smaller exposure group).*

*GNAQ and GNA11 mutations are characteristic of uveal melanoma, which typically lacks BRAF, RAS, and NF1 mutations and carries a poor prognosis, so a crude association between either marker and death is confounded by subtype. The joint model enters both markers and noncutaneous subtype together; the third block reestimates them within cutaneous melanoma only, where noncutaneous subtypes are excluded by design. The Gq-pathway association was not evident after accounting for subtype; the triple-wild-type association is attenuated but persists within cutaneous melanoma (HR, 1.41; 95% CI, 1.06-1.89) and is therefore not entirely explained by subtype.*

### eTable 7. Interaction and Age-Modification Results.

| Estimate | Encoding | N (deaths) | HR (95% CI) | Likelihood-ratio P |
|---|---|---|---|---|
| **Interactions inside the multivariable model, primary endpoint** | | | | |
| Age at sequencing, per decade × burden | TMB-high | 1063 (445) | 1.29 (1.11-1.50) | <.001 |
| Age at sequencing, per decade × burden | Per doubling | 1063 (445) | 1.13 (1.06-1.19) | <.001 |
| Male sex × burden | TMB-high | 1063 (445) | 0.87 (0.59-1.29) | .50 |
| Male sex × burden | Per doubling | 1063 (445) | 1.00 (0.85-1.18) | >.99 |
| Noncutaneous subtype × burden | TMB-high | 1063 (445) | 1.08 (0.46-2.55) | .86 |
| Noncutaneous subtype × burden | Per doubling | 1063 (445) | 1.06 (0.81-1.39) | .69 |
| **Age × burden interaction across secondary endpoints** | | | | |
| Overall survival, from first melanoma diagnosis | TMB-high | 1063 (445) | 1.30 (1.12-1.50) | <.001 |
| Melanoma-specific survival | TMB-high | 702 (160) | 1.43 (1.13-1.81) | .003 |
| Any-cancer-specific survival | TMB-high | 702 (183) | 1.31 (1.05-1.63) | .01 |
| **Burden HR within age quartiles — multivariable covariate set** | | | | |
| Q1 (4-56 y) | TMB-high | 269 (84) | 0.57 (0.35-0.95) | P=.03 |
| Q2 (56-66 y) | TMB-high | 263 (99) | 0.58 (0.38-0.90) | P=.02 |
| Q3 (66-74 y) | TMB-high | 266 (111) | 0.51 (0.33-0.80) | P=.003 |
| Q4 (74-102 y) | TMB-high | 265 (151) | 0.91 (0.58-1.44) | P=.68 |
| Trend across quartiles (1 df) | TMB-high | 1063 (445) | 1.21 (1.03-1.43) | .02 |
| Heterogeneity across quartiles (3 df) | TMB-high | 1063 (445) | — | .04 |
| Q1 (4-56 y) | Per doubling | 269 (84) | 0.68 (0.53-0.87) | P=.002 |
| Q2 (56-66 y) | Per doubling | 263 (99) | 0.75 (0.62-0.90) | P=.002 |
| Q3 (66-74 y) | Per doubling | 266 (111) | 0.79 (0.67-0.95) | P=.01 |
| Q4 (74-102 y) | Per doubling | 265 (151) | 0.87 (0.75-1.00) | P=.046 |
| Trend across quartiles (1 df) | Per doubling | 1063 (445) | — | .06 |
| Heterogeneity across quartiles (3 df) | Per doubling | 1063 (445) | — | .09 |

| Estimate | Encoding | N (deaths) | HR (95% CI) | Likelihood-ratio P |
|---|---|---|---|---|
| **Burden HR within age quartiles — age as a stratification variable, sex and noncutaneous subtype** | | | | |
| Q1 (4-56 y) | TMB-high | 269 (84) | 0.54 (0.33-0.87) | P=.01 |
| Q2 (56-66 y) | TMB-high | 263 (99) | 0.56 (0.37-0.84) | P=.005 |
| Q3 (66-74 y) | TMB-high | 266 (111) | 0.56 (0.38-0.84) | P=.005 |
| Q4 (74-102 y) | TMB-high | 265 (151) | 0.98 (0.66-1.46) | P=.93 |
| One HR common to all four strata | TMB-high | 1063 (445) | 0.65 (0.52-0.81) | — |
| Trend across quartiles (1 df) | TMB-high | 1063 (445) | 1.23 (1.01-1.48) | .03 |
| Heterogeneity across quartiles (3 df) | TMB-high | 1063 (445) | — | .08 |
| **Age functional-form sensitivity, within the multivariable model (N = 1063; 445 deaths)** | | | | |
| Linear, per decade (prespecified) | TMB-high | 1063 (445) | 1.29 (1.11-1.50) | <.001 |
| Age nonlinearity, 4-knot spline vs linear | — | 1063 (445) | — | .009 |
| 4-knot spline age, linear product term | TMB-high | 1063 (445) | 1.23 (1.06-1.44) | .006 |
| 3-knot spline age, linear product term | TMB-high | 1063 (445) | 1.22 (1.05-1.43) | .008 |

*Abbreviations: df, degrees of freedom; HR, hazard ratio; Q, quartile; TMB, tumor mutational burden.*

*The first block shows the 3 exposure-by-covariate product terms available in the multivariable model, each fitted within that model and tested by likelihood ratio against the same model without the product term; this set is complete rather than selected, and age was designated in advance as the interaction of primary interest. Cause-specific end points are administratively censored at December 7, 2020 (eTable 3). The quartile blocks are given under two specifications: the first refits sex and noncutaneous subtype within each stratum; the second gives each quartile its own baseline hazard, with sex and noncutaneous subtype as the other terms. All blocks use the analytic cohort (N = 1063). Each quartile block carries a 1-degree-of-freedom test for linear trend alongside the conventional unstructured heterogeneity test; the trend test accounts for the ordering of the quartiles, which the unstructured test ignores. The final block refits the age interaction under alternative specifications of age and includes a test of nonlinearity in the age main effect.*

**eFigure 1. Cohort Flow Diagram.**

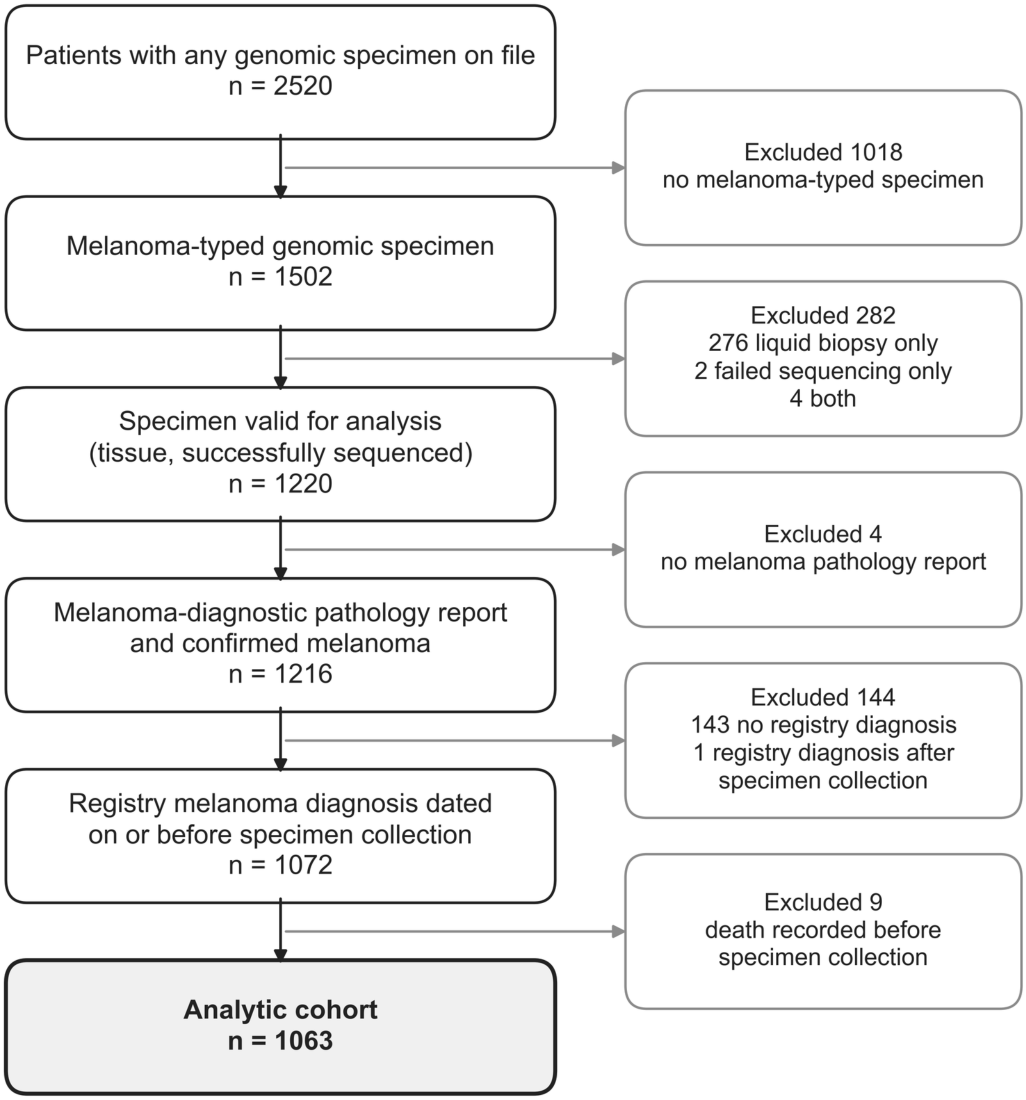


*The bold box indicates the analytic cohort, in which all analyses were conducted. Eligibility was assessed at the patient rather than the specimen level, and one representative tissue specimen was selected per patient.*

**eFigure 2. Cumulative Incidence of Melanoma Death by Tumor Mutational Burden Status.**

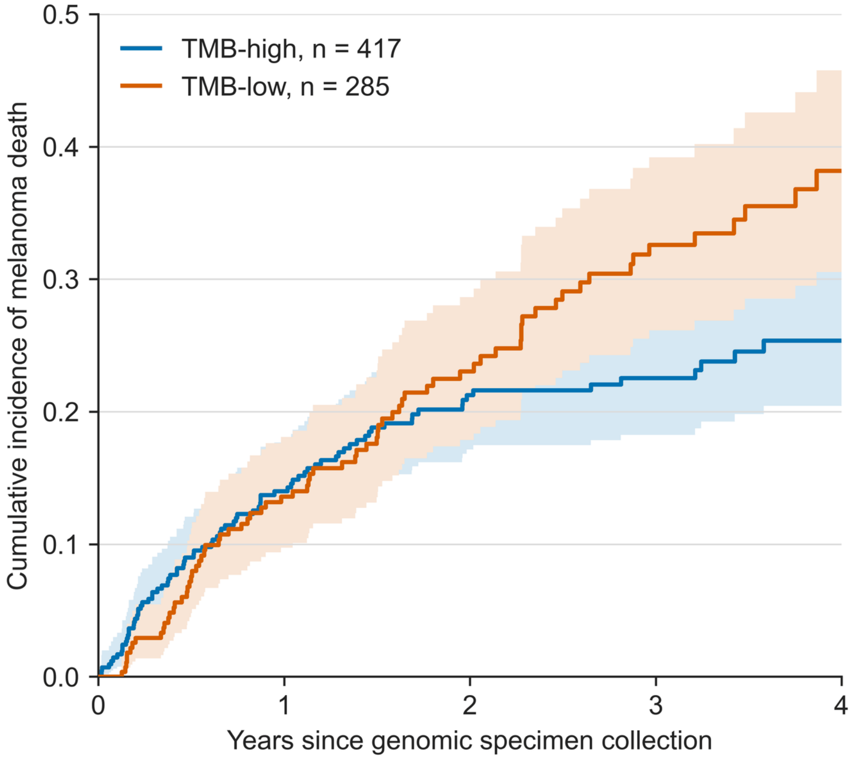


| **No. at risk** | | | | | |
|---|---|---|---|---|---|
| TMB-high | 417 | 289 | 201 | 138 | 70 |
| TMB-low | 285 | 201 | 126 | 81 | 36 |

*Death from another cause is treated as a competing event (Aalen-Johansen estimator). Follow-up is administratively censored at the National Death Index cause-of-death linkage cutoff (December 7, 2020), because cause was ascertained for 97.2% of deaths on or before that date and none after it, so only the 702 patients sequenced before that date contribute; without that censoring the cause-specific hazard is estimated as zero past the cutoff and absolute risk is understated. Shaded areas are 95% confidence bands. Curves are truncated at 4 years, when at least 10% of each group remained at risk. The corresponding cause-specific hazard ratios are in eTable 4, and their sensitivity to the handling of the cutoff is in eTable 3. TMB indicates tumor mutational burden; TMB-high, ≥10 mutations per megabase.*

### eFigure 3. Delayed-Entry (Left-Truncation) Sensitivity Analysis.

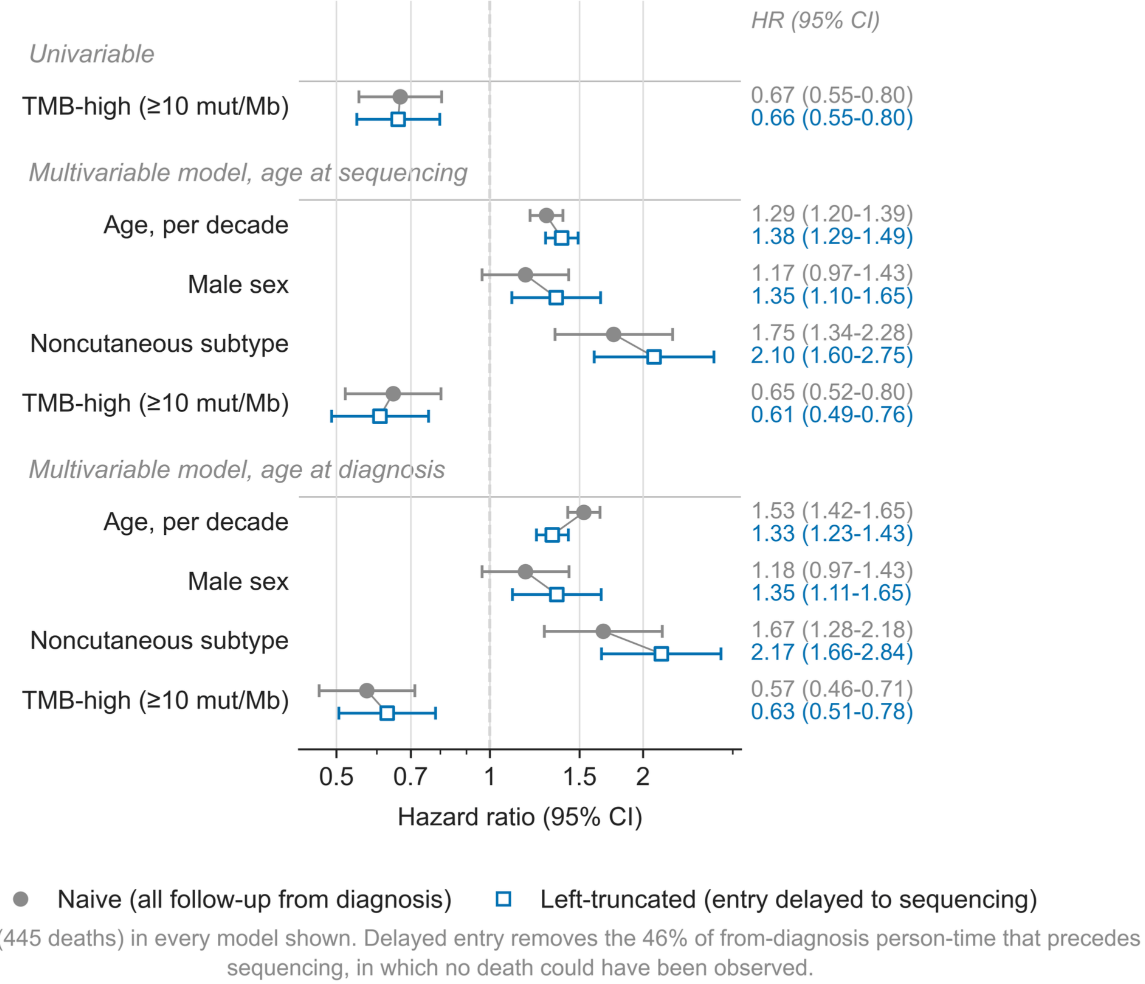


*Hazard ratios estimated naively, using all follow-up from diagnosis, compared with the same models under delayed entry at the date of genomic specimen collection, so that no patient contributes person-time during which a death could not have been observed. All models include the same 1063 patients (445 deaths). The univariable model includes TMB-high only; the multivariable models include age (at sequencing or at diagnosis), sex, noncutaneous subtype, and TMB-high. The burden estimate is essentially unchanged, indicating that immortal time bias does not account for the association. HR indicates hazard ratio; mut/Mb, mutations per megabase; TMB, tumor mutational burden; TMB-high, ≥10 mut/Mb.*

### eFigure 4. The Age × TMB-High Interaction Under Alternative Specifications of Age.

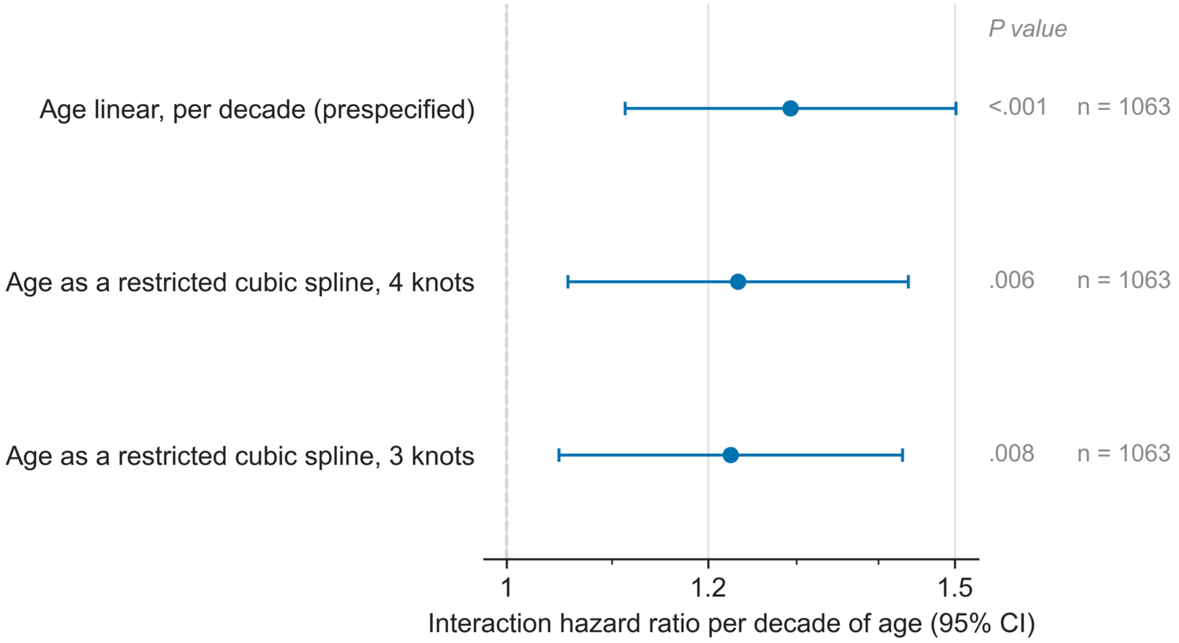


Nonlinearity of the main effect of age: 4-knot *P* = .009; 3-knot *P* = .009
With age as a stratification variable, the common TMB-high hazard ratio was 0.65 (95% CI, 0.52-0.81)
(a main effect, not an interaction, so it is not plotted).

*Because a product term can absorb curvature left unmodeled in a main effect and TMB-high patients were on average 8 years older than TMB-low patients, the interaction was refit with alternative specifications of age within the prespecified multivariable model. The first row enters age as linear (the prespecified specification); the second and third model the main effect of age as a restricted cubic spline (4 and 3 knots) while keeping the product term linear. All rows use the analytic cohort (n = 1063; 445 deaths). Only specifications whose estimate is an interaction hazard ratio per decade are plotted, so that every marker is on one scale. P values are from likelihood ratio tests (LRTs) against the same model without the interaction term. TMB indicates tumor mutational burden; TMB-high, ≥10 mutations per megabase.*

**eFigure 5. Modification of the Tumor Mutational Burden Association by Age, With Burden Modeled Continuously.**

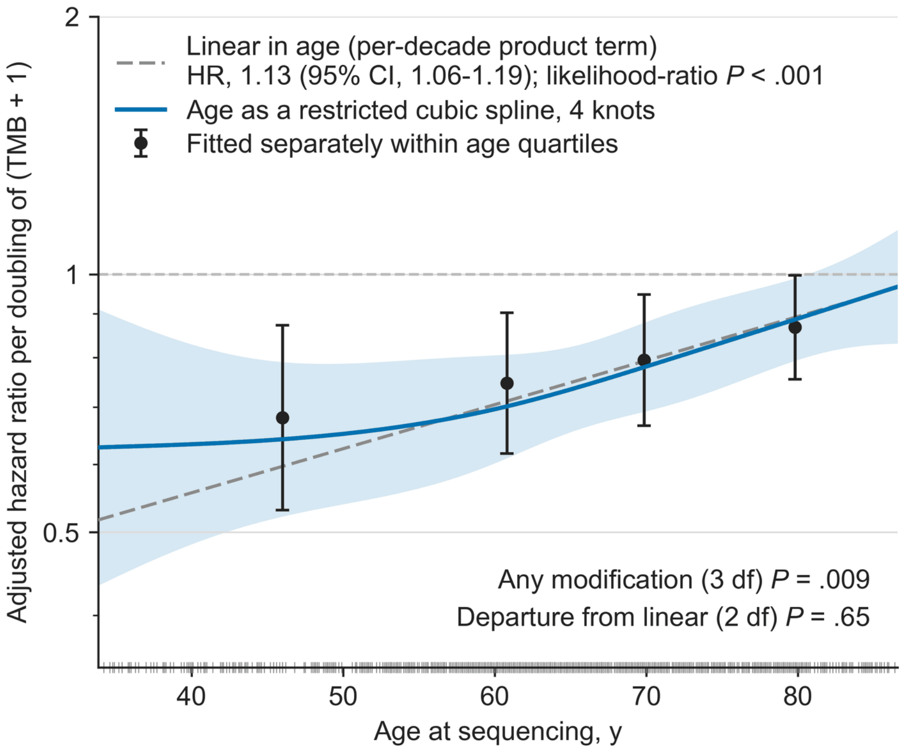


*The construction is that of Figure 2, with the exposure entered as log2(TMB + 1), a hazard ratio per doubling of burden, in place of the 10 mut/Mb dichotomy; model, sample (n = 1063; 445 deaths), covariates, knot placement, confidence-interval method and plotting range are otherwise identical. Modification by age was also present under this encoding (3-df likelihood ratio test, P = .009) and did not depart from linearity in age (2-df test, P = .65), so the age dependence reported in the main text is not a consequence of dichotomizing burden at 10 mut/Mb. Circles are the per-doubling hazard ratio fitted separately within quartiles of age, shown for orientation only, as in Figure 2. HR indicates hazard ratio; TMB, tumor mutational burden.*
